# Social Determinants of Cardiovascular Disease in US Women with and without Breast Cancer

**DOI:** 10.1101/2024.06.24.24309445

**Authors:** Elena Wadden, Vidhushei Yogeswaran, Roberta M Ray, Alexi Vasbinder, Aladdin H Shadyab, Qian Xiao, Phyllis A Richey, Nazmus Saquib, Yangbo Sun, Su Yon Jung, Margaret S. Pichardo, JoAnn E Manson, Garnet Anderson, Michael Simon, Marcia Stefanick, Kerryn Reding, Ana Barac, Richard K Cheng

**Author notes:** **Corresponding Author:** Richard K Cheng MD MS 1959 NE Pacific Street, Health Sciences Building, Suite #A506D Box 356422 Seattle, WA 98195, or. **Declarations of interest:** none. **Funding:** The WHI program is funded by National Heart, Lung, and Blood Institute, National Institutes of Health, U.S. Department of Health and Human Services through contracts, HHSN268201600018C, HHSN268201600001C, HHSN268201600002C, HHSN268201600003C, and HHSN268201600004C. The authors have reported that they have no relationships relevant to the contents of this paper to disclose.

## Abstract

**Background:** Social determinants of health (SDOH) may be related to CV risk in women with and without breast cancer (BC).

**Methods:** In 153,401 women without prevalent CV disease in the Women’s Health Initiative (WHI), SDOH factors included geographic region, rurality, insurance status, and household income. Cox proportional hazard models were used to assess associations between SDOH factors and a composite CV outcome (myocardial infarction, stroke, hospitalization for heart failure, or CV death). Effect modification by BC was assessed between BC status and SDOH.

**Results:** The final cohort included 10,954 (mean age 62±7 years) women who subsequently developed BC and 142,144 (mean age 63 ± 7) women without BC. During a median follow-up of 13 years, 18,148 women had an incident composite CV outcome. Rurality, low household income, and non-private insurance were associated with an increased risk of composite CV outcome and mortality in both women with and without BC. There was no effect modification by BC status for associations with rurality, insurance, or household income. By geographic region, there was no difference in composite CV outcome in women without BC. However, there was a significant interaction by BC status for geographic region and the composite endpoint (p=0.009). Women with BC in the South had a lower risk of the composite outcome (HR 0.68, 95% CI 0.54-0.87).

**Conclusions:** SDOH are associated with an increased risk of CV events and mortality among women. There were no differential associations based on BC status, suggesting that women with BC face similar vulnerabilities related to SDOH.

## Introduction

Cardiovascular (CV) events and breast cancer (BC) are major causes of disease in postmenopausal women, sharing risk factors and underlying pathophysiologic mechanisms.^1^ In the United States, CV disease is the leading cause of death in women. Women with BC face increased CV risks due to cardiometabolic changes of malignancy, adverse effects of cancer therapeutics, and indirect treatment effects.^2^ This elevated CV risk in postmenopausal women with BC is most pronounced several years after BC diagnosis.^3^

Disparities persist in CV outcomes despite advancements in healthcare, often driven by complex interactions with social determinants of health (SDOH).^2,4^ SDOH, as defined by Centers for Disease Control and Prevention, encompass the conditions under which individuals are born, grow, work, live, and age, and the wider set of forces and systems shaping the conditions of daily life.”^5^ In the general population, CV outcomes exhibit significant variation by socioeconomic status, region of residence, and rurality.^6–8^ In the United States, regional variability in insurance coverage further exacerbate these differences, leading to variability in access and ultimately outcomes.^9^ For women in particular, prior research has highlighted an increased CV risk associated with lower income, rural residence, and lower educational attainment.^10–12^

Despite our understanding of CV and BC risk, there is a critical gap in our knowledge on how SDOH and regional variations affect CV risk in women with BC.^13^ While both women with and without BC have a high burden of CV disease, the extent to which BC status modifies the risk of CV disease remains unanswered. Therefore, we sought to better characterize the associations of SDOH and geographic regions with CV disease outcomes and determine whether BC status modifies the risk of CV disease. In this study, we utilize longitudinal data from the Women’s Health Initiative (WHI) to evaluate the association of geographic differences and SDOH with CV outcomes in both women with and without BC. Understanding associations between SDOH, geographic regions, and CV outcomes may better identify women at higher CV risk for targeted clinical and public health initiatives.

## Methods

### Study Population

The WHI is a longitudinal study of postmenopausal women aged 50 to 79 years who were recruited from 40 US clinical centers between October 1, 1993, and December 31, 1998, with ongoing, longitudinal follow-up.^14–16^ We included women who participated in either the WHI Clinical Trial (n=68,132) or enrolled in an observational study (n=93,676). Participants were followed through March 2005 and had the option to continue follow up in two extension studies through 2022.^17,18^ The institutional review boards from all WHI-affiliated institutions approved this study. All participants provided informed consent. During Extension Study II, cardiovascular outcomes were adjudicated in a subset of participants, referred to as the Medical Records Cohort (MRC n=22,316), which consisted of all women who participated in the hormone trial component of the original clinical trial, as well as all African American and Hispanic women from any study component. The remaining participants comprised the Self-Report Cohort (SRC n=71,251). Cancer was adjudicated in both the MRC and the SRC. The present analysis excluded women with self-reported history of myocardial infarction, coronary revascularization, stroke, or heart failure at baseline (n=7,757). We further excluded women with no follow-up after baseline (n=650) (Figure I).

### Outcomes & Adjudication process

The primary outcome was a composite of incident CV events (non-fatal myocardial infarction, stroke, and hospitalization for heart failure) and CVD death (excluding fatal pulmonary embolism). Cause of death was determined by trained physician adjudicators for all reports of a participant death through Extension Study I.^19^ In Extension Study II, cause of death was adjudicated only in the Medical Record Cohort. Deaths in the Self-Report Cohort were identified, and cause of death determined by annual linkage to the National Death Index.^19^ We also considered CVD mortality, breast cancer mortality, and all-cause mortality, separately, as secondary outcomes. Incident CV events and breast cancer were initially ascertained by self-report, then documented by medical record review. CVD cases were evaluated by local trained WHI-physician adjudicators for evaluation and classification, with locally adjudicated cases sent to WHI Clinical Coordinating Center for central adjudication.^17^ Events were defined prospectively using established criteria. Using methods from the National Cancer Institute Surveillance, Epidemiology, and End Results guidelines, breast cancer cases were centrally coded.^20^

### Primary Exposures

Exposures of interest included region in the United States, rural-urban commuting area codes (RUCA), insurance status, and household income. These were obtained at the time of WHI study enrollment.

Geographic region of residence was divided into the 4 US Census Bureau regions 1) Northeast included Pennsylvania, New Jersey, New York, Connecticut, Massachusetts, Vermont, New Hampshire, and Maine. 2) South included Texas, Oklahoma, Louisiana, Arkansas, Mississippi, Alabama, Tennessee, Kentucky, West Virginia, Virginia, Maryland, Delaware, North Carolina, South Carolina, Georgia, and Florida. 3) Midwest included Kansas, Nebraska, South Dakota, North Dakota, Missouri, Iowa, Minnesota, Illinois, Wisconsin, Indiana, Ohio, and Michigan. 4) West included Montana, Idaho, Wyoming, Nevada, Utah, Colorado, Arizona, New Mexico, California, Oregon, Washington, Alaska, and Hawaii.

RUCA codes were used to classify United States census tracts into 1 of 10 main categories based on rural-urban core and extent of commuting. Based on methodology used from previously published WHI studies looking at the rural-urban classification and based on the distribution of WHI participants across the country, we classified women in our study into one of three classes at time of WHI study entry: 1) urban area, 2) large rural city/town, and 3) small rural town or isolated small rural town.^21^

Insurance status was divided into participants with Medicare or Medicaid; HMO or other private; combination of public and private; military, VA, or other insurance only; no insurance; or unknown. Household income was evaluated in four categories, similar to previously published studies in the WHI: 1) < $20,000, 2) $20,000-$34,999, 3) $35,000-$49,999, and 4) ≥$50,000.^22^

### Covariates

Baseline characteristics were obtained at WHI study enrollment as previously described.^17,23^ In addition to the primary exposures listed above, self-reported information on demographics (race and ethnicity, age at enrollment, and education were collected), lifestyle factors (smoking history and physical activity), comorbidities (history of hypertension, diabetes, and hyperlipidemia), and family medical history (myocardial infarction and stroke) were collected. Smoking history was self-defined as either current, former, or never smoker. Physical activity was measured as total metabolic equivalent (MET) hours per week calculated from self-report questionnaires. Body mass index was collected through in-person clinic visits at enrollment and was calculated as weight (kg) divided by the square of the body height (m^2^). *Statistical Analysis*

Baseline characteristics were stratified by RUCA code and geographic region and described as mean values ± standard deviation for continuous variables and as frequencies and percentages for categorical variables. We used Cox proportional hazard regression models to assess the associations between SDOH and outcomes in stepwise models. Model 1 included age at enrollment and all SDOH variables (i.e., geographic region of residence, RUCA, insurance status, and household income). Model 2 included the variables in model 1 in addition to physical activity, smoking status, diabetes, hypertension, hyperlipidemia, and family history of myocardial infarction and stroke. Lastly, model 3 included all variables in model 2 plus adjustment for race, ethnicity, and body mass index. Associations were presented as hazard ratios (HRs) and 95% confidence intervals. Invasive BC during follow-up was included in all models as a time-varying covariate in order to identify BC relative to the timing of the CV outcome. Effect modification by BC status (BC vs non-BC) was assessed by including interaction terms between BC status and each SDOH variables, in separate fully adjusted models (i.e., model 3). HRs were estimated from the interaction models and BC mortality was modeled as competing risk in the Cox PH analyses. The time scale was defined as time since the enrollment in the WHI. For the primary endpoint, follow-up was censored in SRC participants at the end of Extension I when adjudication of non-fatal CV events ended. Follow-up for MRC participants continued to the last documented follow-up time or death from other cause, as of February 19, 2022. For the secondary endpoint of CV death, all participants without the endpoint were censored at last documented follow-up time or death from other cause, as of February 19, 2022. We conducted additional analyses for BC specific mortality, death from other causes, and all-cause death separately.

Final models were conducted on participants with complete covariate and exposure data. All statistical analyses were 2-sided and completed using SAS version 9.4 software (SAS Institute Inc., Cary, NC, USA). P values of <0.05 were considered statistically significant.

## Results

### Baseline characteristics of study cohort

In our final cohort of 153,401 participants, there were 10,954 (mean age at baseline 62±7 years) women who subsequently developed incident invasive BC during the study period. From enrollment to the end of their follow-up, 94% of women resided in the same broad geographic region since baseline collection, and 84% of women had no RUCA category change. Among women who developed BC, the mean (±SD) age at BC diagnosis was 73±8 years, 90% were White, 6% Black, 2% Asian/Pacific Islander, and 2% more than one race/unknown. There was a higher prevalence of women with BC living in small rural and large rural areas in the Midwest (52% and 37.6% respectively) compared to the Northeast (Table I). Compared to those in urban areas, women with BC in large rural and small rural areas were more likely to be uninsured (3% versus 5-7%), have a high school diploma or less (17% versus 27-29%), and have an income level <$20,000 (10% versus 14-15%). When stratified by geographic region, there were no differences in baseline demographics, socioeconomic status, or health behaviors (Table 2).

**Table 1.**
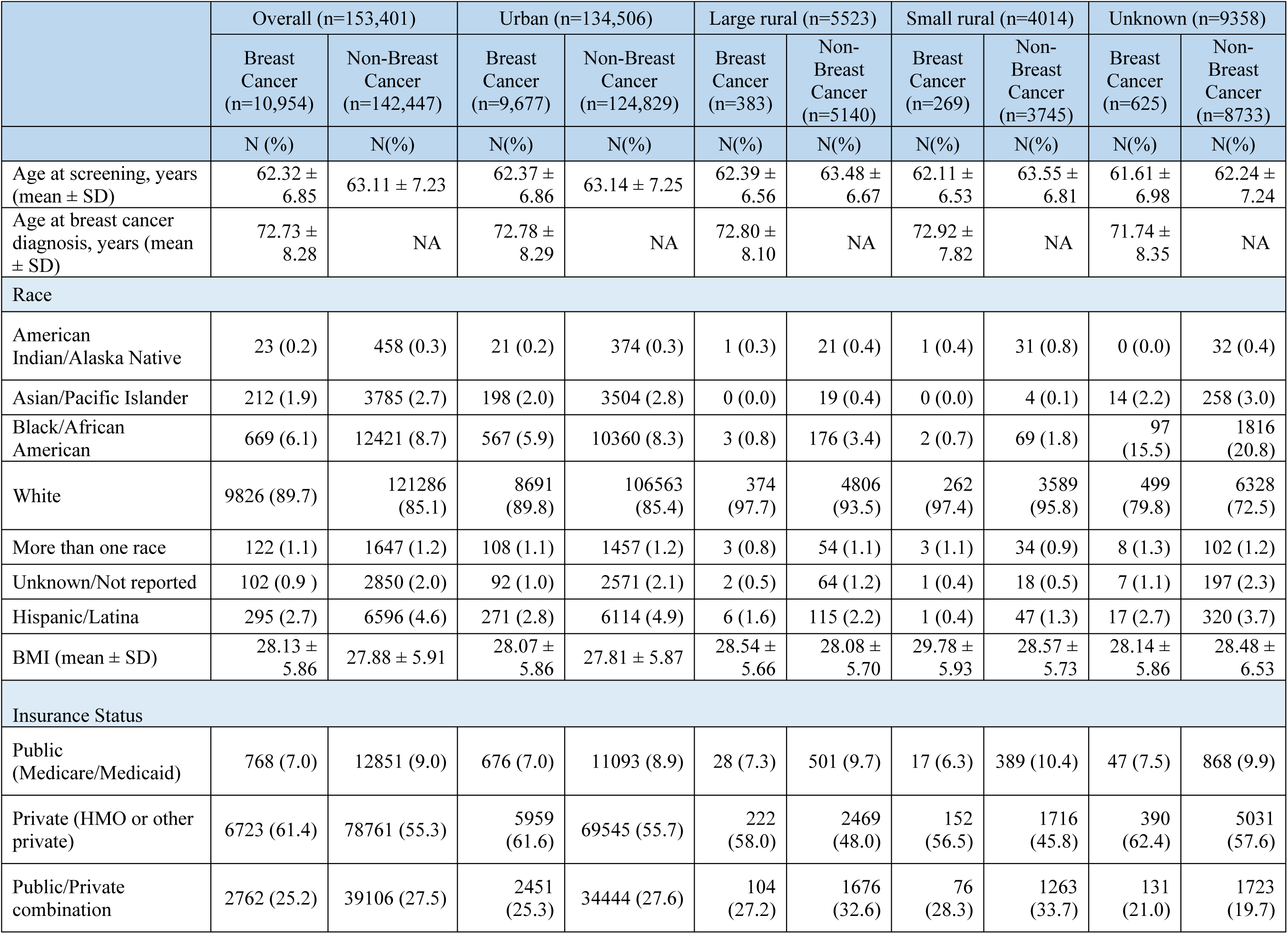
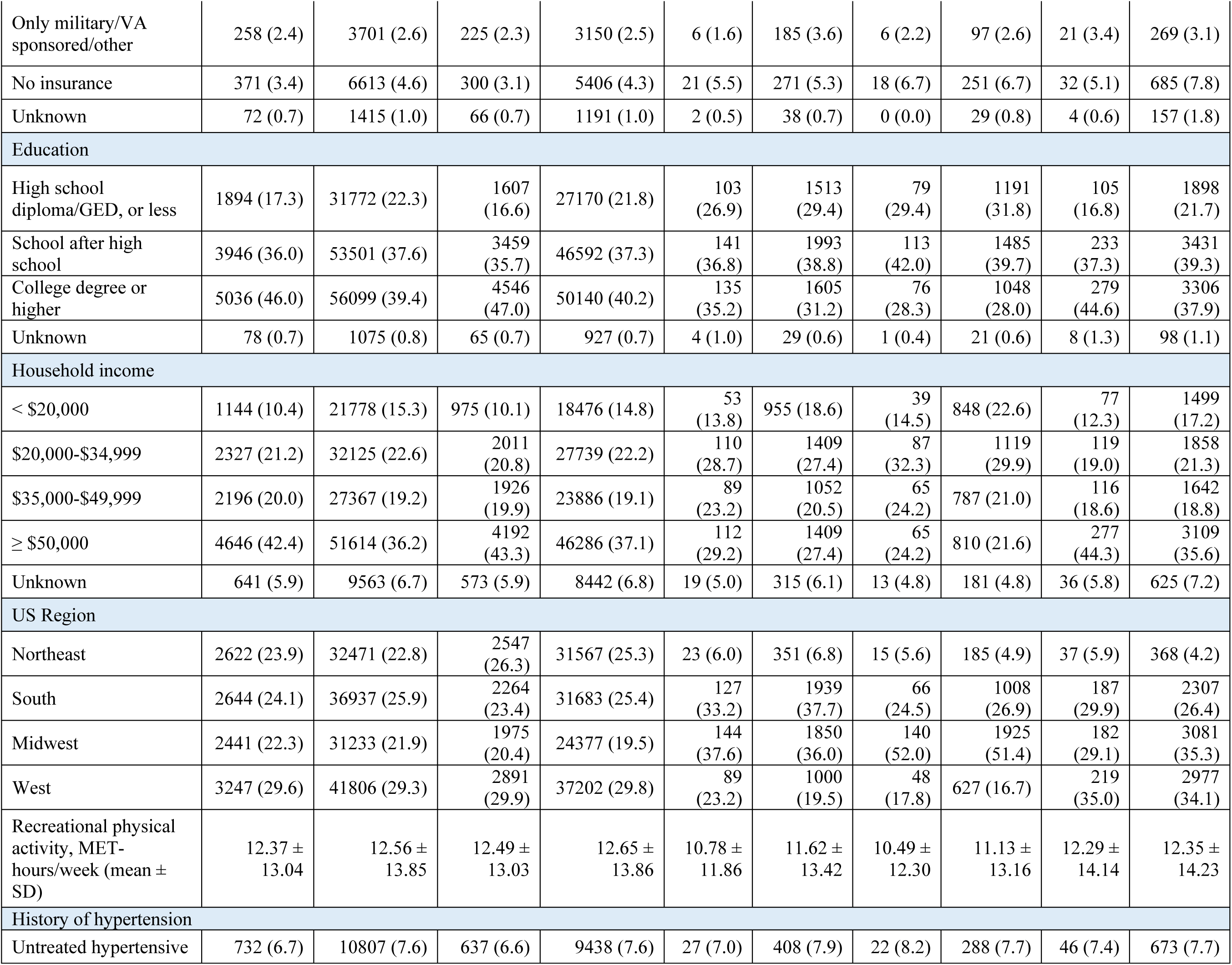
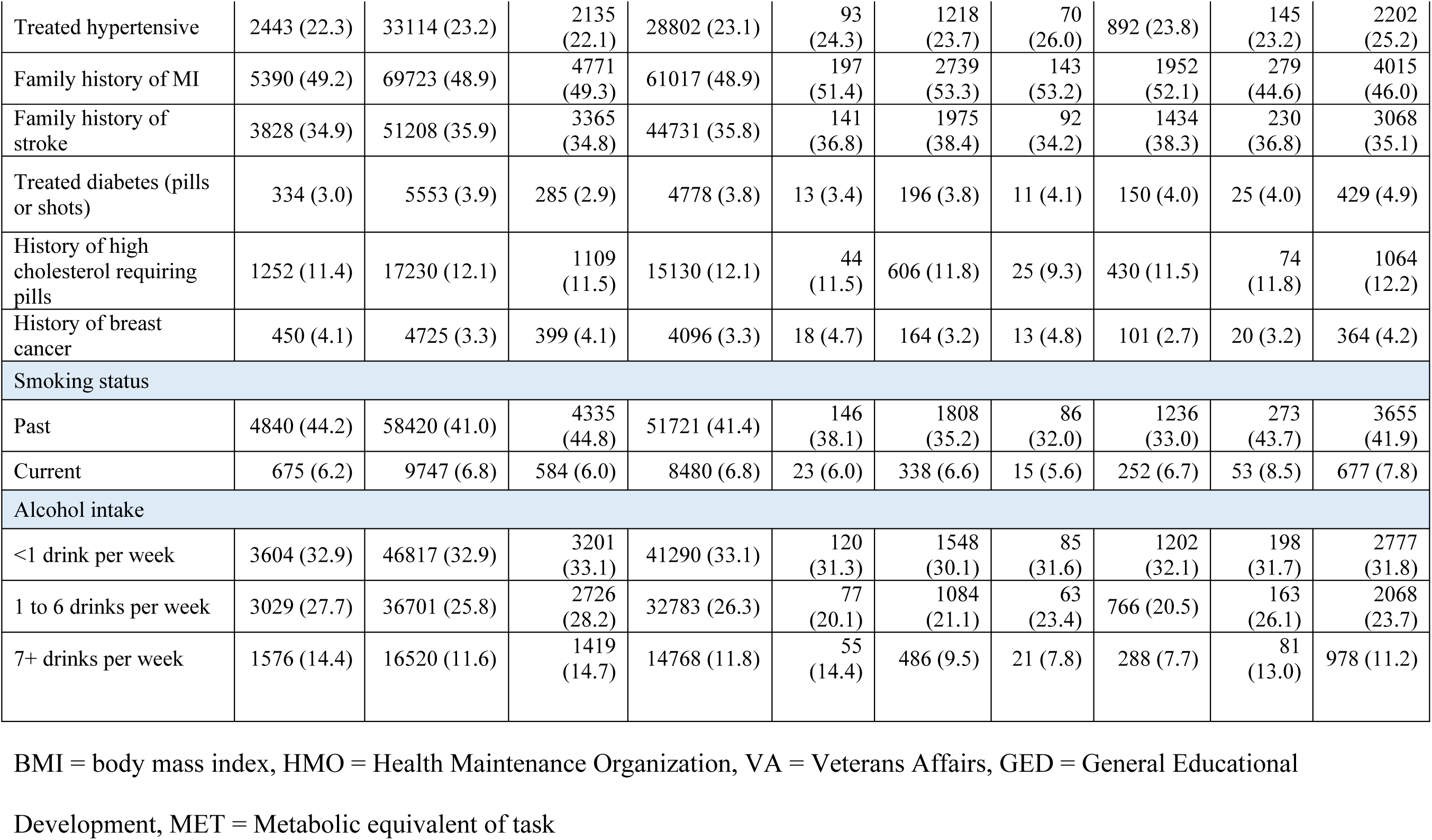
Baseline characteristics by RUCA.

**Table 2.** Baseline characteristics of WHI cohort by geographic region.

|  | Overall (n=153,401) |  | Northeast (n=35,093) |  | South (n=39,581) |  | Midwest (n=33,674) |  | West (n=45,053) |  |
| --- | --- | --- | --- | --- | --- | --- | --- | --- | --- | --- |
|  | Breast Cancer<br>(n=10,954) | Non-Breast Cancer<br>(n=142,447) | Breast Cancer<br>(n=2,622) | Non-Breast Cancer<br>(n=32,471) | Breast Cancer<br>(n=2,644) | Non-Breast Cancer<br>(n=36,937) | Breast Cancer<br>(n=2,441) | Non-Breast Cancer<br>(n=31,233) | Breast Cancer<br>(n=3,247) | Non-Breast Cancer<br>(n=41,806) |
|  | N(%) | N(%) | N(%) | N(%) | N(%) | N(%) | N(%) | N(%) | N(%) | N(%) |
| Age at screening, years (mean ± SD) | 62.32 ± 6.85 | 63.11 ± 7.23 | 62.52 ± 6.56 | 63.37 ± 6.94 | 61.44 ± 6.76 | 62.25 ± 7.24 | 62.21 ± 6.74 | 63.06 ± 7.03 | 62.96 ± 7.15 | 63.71 ± 7.49 |
| Age at breast cancer diagnosis, years (mean ± SD) | 72.73 ± 8.28 | | 73.46 ± 8.14 | | 71.76 ± 8.24 | | 72.59 ± 8.26 | | 73.01 ± 8.35 | |
| Race |  |  |  |  |  |  |  |  |  |  |
| American Indian/Alaska Native | 23 (0.2) | 458 (0.3) | 4 (0.2) | 43 (0.1) | 5 (0.2) | 117 (0.3) | 6 (0.2) | 36 (0.1) | 8 (0.2) | 262 (0.6) |
| Asian/Pacific Islander | 212 (1.9) | 3785 (2.7) | 9 (0.3) | 195 (0.6) | 9 (0.3) | 242 (0.7) | 10 (0.4) | 161 (0.5) | 184 (5.7) | 3187 (7.6) |
| Black/African American | 669 (6.1) | 12421 (8.7) | 110 (4.2) | 2132 (6.6) | 343 (13.0) | 5936 (16.1) | 150 (6.1) | 2961 (9.5) | 66 (2.0) | 1392 (3.3) |
| White | 9826 (89.7) | 121286 (85.1) | 2463 (93.9) | 29429 (90.6) | 2245 (84.9) | 29524 (79.9) | 2253 (92.3) | 27631 (88.5) | 2865 (88.2) | 34702 (83.0) |
| More than one race | 122 (1.1) | 1647 (1.2) | 19 (0.7) | 251 (0.8) | 24 (0.9) | 334 (0.9) | 15 (0.6) | 239 (0.8) | 64 (2.0) | 823 (2.0) |
| Unknown/Not reported | 102 (0.9) | 2850 (2.0) | 17 (0.6) | 421 (1.3) | 18 (0.7) | 784 (2.1) | 7 (0.3) | 205 (0.7) | 60 (1.8) | 1440 (3.4) |
| Hispanic/Latina | 295 (2.7) | 6596 (4.6) | 38 (1.4) | 870 (2.7) | 101 (3.8) | 2592 (7.0) | 12 (0.5) | 288 (0.9) | 144 (4.4) | 2846 (6.8) |
| BMI (mean ± SD) | 28.13 ± 5.86 | 27.88 ± 5.91 | 28.32 ± 5.82 | 27.96 ± 5.82 | 28.06 ± 5.85 | 27.96 ± 6.05 | 28.54 ± 5.92 | 28.33 ± 5.99 | 27.74 ± 5.83 | 27.41 ± 5.75 |
| BMI ≥30 | 3403 (31.1) | 41646 (29.2) | 866 (33.0) | 9631 (29.7) | 796 (30.1) | 11068 (30.0) | 836 (34.2) | 10031 (32.1) | 905 (27.9) | 10916 (26.1) |
| Insurance Status |  |  |  |  |  |  |  |  |  |  |
| Public (Medicare/Medicaid) | 768 (7.0) | 12851 (9.0) | 210 (8.0) | 3323 (10.2) | 202 (7.6) | 3765 (10.2) | 146 (6.0) | 2496 (8.0) | 210 (6.5) | 3267 (7.8) |
| Private (HMO or other private) | 6723 (61.4) | 78761 (55.3) | 1638 (62.5) | 18368 (56.6) | 1569 (59.3) | 19431 (52.6) | 1490 (61.0) | 16885 (54.1) | 2026 (62.4) | 24077 (57.6) |
| Public/Private combination | 2762 (25.2) | 39106 (27.5) | 654 (24.9) | 9124 (28.1) | 592 (22.4) | 8912 (24.1) | 698 (28.6) | 9980 (32.0) | 818 (25.2) | 11090 (26.5) |
| Only military/<br>VA sponsored/<br>other | 258 (2.4) | 3701 (2.6) | 36 (1.4) | 473 (1.5) | 119 (4.5) | 1686 (4.6) | 31 (1.3) | 482 (1.5) | 72 (2.2) | 1060 (2.5) |
| No insurance | 371 (3.4) | 6613 (4.6) | 68 (2.6) | 931 (2.9) | 137 (5.2) | 2527 (6.8) | 60 (2.5) | 1156 (3.7) | 106 (3.3) | 1999 (4.8) |
| Unknown | 72 (0.7) | 1415 (1.0) | 16 (0.6) | 252 (0.8) | 25 (0.9) | 616 (1.7) | 16 (0.7) | 234 (0.7) | 15 (0.5) | 313 (0.7) |
| Education |  |  |  |  |  |  |  |  |  |  |
| High school<br>diploma/GED, or<br>less | 1894 (17.3) | 31772 (22.3) | 511 (19.5) | 8291 (25.5) | 461 (17.4) | 8411 (22.8) | 475 (19.5) | 7472 (23.9) | 447 (13.8) | 7598 (18.2) |
| School after high<br>school | 3946 (36.0) | 53501 (37.6) | 847 (32.3) | 10910<br>(33.6) | 970 (36.7) | 13948<br>(37.8) | 865 (35.4) | 11155<br>(35.7) | 1264<br>(38.9) | 17488<br>(41.8) |
| College degree<br>or higher | 5036 (46.0) | 56099 (39.4) | 1245<br>(47.5) | 13057<br>(40.2) | 1191<br>(45.0) | 14209<br>(38.5) | 1083<br>(44.4) | 12376<br>(39.6) | 1517<br>(46.7) | 16457<br>(39.4) |
| Unknown | 78 (0.7) | 1075 (0.8) | 19 (0.7) | 213 (0.7) | 22 (0.8) | 369 (1.0) | 18 (0.7) | 230 (0.7) | 19 (0.6) | 263 (0.6) |
| Household income |  |  |  |  |  |  |  |  |  |  |
| < \$20,000 | 1144 (10.4) | 21778 (15.3) | 282 (10.8) | 4687 (14.4) | 305 (11.5) | 6272 (17.0) | 240 (9.8) | 4558 (14.6) | 317 (9.8) | 6261 (15.0) |
| \$20,000-\$34,999 | 2327 (21.2) | 32125 (22.6) | 504 (19.2) | 7183 (22.1) | 519 (19.6) | 7843 (21.2) | 604 (24.7) | 7548 (24.2) | 700 (21.6) | 9551 (22.8) |
| \$35,000-\$49,999 | 2196 (20.0) | 27367 (19.2) | 492 (18.8) | 6080 (18.7) | 513 (19.4) | 6911 (18.7) | 512 (21.0) | 6305 (20.2) | 679 (20.9) | 8071 (19.3) |
| ≥ \$50,000 | 4646 (42.4) | 51614 (36.2) | 1155<br>(44.1) | 12146<br>(37.4) | 1151<br>(43.5) | 13128<br>(35.5) | 946 (38.8) | 10736<br>(34.4) | 1394<br>(42.9) | 15604<br>(37.3) |
| Unknown | 641 (5.9) | 9563 (6.7) | 189 (7.2) | 2375 (7.3) | 156 (5.9) | 2783 (7.5) | 139 (5.7) | 2086 (6.7) | 157 (4.8) | 2319 (5.5) |
| Rural-Urban Residence (RUCA) |  |  |  |  |  |  |  |  |  |  |
| Urban | 9677 (88.3) | 124829<br>(87.6) | 2547<br>(97.1) | 31567<br>(97.2) | 2264<br>(85.6) | 31683<br>(85.8) | 1975<br>(80.9) | 24377<br>(78.0) | 2891<br>(89.0) | 37202<br>(89.0) |
| Large rural | 383 (3.5) | 5140 (3.6) | 23 (0.9) | 351 (1.1) | 127 (4.8) | 1939 (5.2) | 144 (5.9) | 1850 (5.9) | 89 (2.7) | 1000 (2.4) |
| Small rural | 134 (1.2) | 1996 (1.4) | 5 (0.2) | 90 (0.3) | 30 (1.1) | 580 (1.6) | 78 (3.2) | 1048 (3.4) | 21 (0.6) | 278 (0.7) |
| Isolated small<br>rural | 135 (1.2) | 1749 (1.2) | 10 (0.4) | 95 (0.3) | 36 (1.4) | 428 (1.2) | 62 (2.5) | 877 (2.8) | 27 (0.8) | 349 (0.8) |
| Missing<br>(no address) | 625 (5.7) | 8733 (6.1) | 37 (1.4) | 368 (1.1) | 187 (7.1) | 2307 (6.2) | 182 (7.5) | 3081 (9.9) | 219 (6.7) | 2977 (7.1) |
| Recreational<br>physical activity,<br>MET-hours/week<br>(mean ± SD) | 12.37 ±<br>13.04 | 12.56 ±<br>13.85 | 11.99 ±<br>12.66 | 12.21 ±<br>13.63 | 11.99 ±<br>13.37 | 11.66 ±<br>13.61 | 11.70 ±<br>11.87 | 12.25 ±<br>13.28 | 13.48 ±<br>13.83 | 13.84 ±<br>14.55 |
| History of hypertension |  |  |  |  |  |  |  |  |  |  |
| Untreated<br>hypertensive | 732 (6.7) | 10807 (7.6) | 169 (6.4) | 2501 (7.7) | 171 (6.5) | 2702 (7.3) | 150 (6.1) | 2244 (7.2) | 242 (7.5) | 3360 (8.0) |
| Treated hypertensive | 2443 (22.3) | 33114 (23.2) | 622 (23.7) | 7537 (23.2) | 593 (22.4) | 8863 (24.0) | 577 (23.6) | 7594 (24.3) | 651 (20.0) | 9120 (21.8) |
| Family history of MI | 5390 (49.2) | 69723 (48.9) | 1333 (50.8) | 16596 (51.1) | 1292 (48.9) | 17952 (48.6) | 1255 (51.4) | 15612 (50.0) | 1510 (46.5) | 19563 (46.8) |
| Family history of stroke | 3828 (34.9) | 51208 (35.9) | 885 (33.8) | 11256 (34.7) | 939 (35.5) | 13484 (36.5) | 909 (37.2) | 11526 (36.9) | 1095 (33.7) | 14942 (35.7) |
| Treated diabetes (pills or shots) | 334 (3.0) | 5553 (3.9) | 78 (3.0) | 1269 (3.9) | 95 (3.6) | 1623 (4.4) | 77 (3.2) | 1173 (3.8) | 84 (2.6) | 1488 (3.6) |
| History of high cholesterol requiring pills | 1252 (11.4) | 17230 (12.1) | 322 (12.3) | 4069 (12.5) | 321 (12.1) | 4664 (12.6) | 268 (11.0) | 3645 (11.7) | 341 (10.5) | 4852 (11.6) |
| History of breast cancer | 450 (4.1) | 4725 (3.3) | 108 (4.1) | 889 (2.7) | 121 (4.6) | 1517 (4.1) | 120 (4.9) | 1181 (3.8) | 101 (3.1) | 1138 (2.7) |
| Smoking status |  |  |  |  |  |  |  |  |  |  |
| Past | 4840 (44.2) | 58420 (41.0) | 1293 (49.3) | 14662 (45.2) | 1093 (41.3) | 13993 (37.9) | 1009 (41.3) | 12466 (39.9) | 1445 (44.5) | 17299 (41.4) |
| Current | 675 (6.2) | 9747 (6.8) | 136 (5.2) | 2289 (7.0) | 170 (6.4) | 2705 (7.3) | 159 (6.5) | 2171 (7.0) | 210 (6.5) | 2582 (6.2) |
| Alcohol intake |  |  |  |  |  |  |  |  |  |  |
| <1 drink per week | 3604 (32.9) | 46817 (32.9) | 903 (34.4) | 11612 (35.8) | 814 (30.8) | 10886 (29.5) | 884 (36.2) | 11245 (36.0) | 1003 (30.9) | 13074 (31.3) |
| 1 to 6 drinks per week | 3029 (27.7) | 36701 (25.8) | 799 (30.5) | 9246 (28.5) | 614 (23.2) | 7971 (21.6) | 689 (28.2) | 8406 (26.9) | 927 (28.5) | 11078 (26.5) |
| 7+ drinks per week | 1576 (14.4) | 16520 (11.6) | 405 (15.4) | 3978 (12.3) | 345 (13.0) | 3662 (9.9) | 303 (12.4) | 3097 (9.9) | 523 (16.1) | 5783 (13.8) |
| Missing | 74 (0.7) | 1073 (0.8) | 22 (0.8) | 200 (0.6) | 21 (0.8) | 420 (1.1) | 9 (0.4) | 169 (0.5) | 22 (0.7) | 284 (0.7) |

- 1 BMI = body mass index, HMO = Health Maintenance Organization, VA = Veterans Affairs, GED = General Educational Development, MET = Metabolic equivalent of task

### Risk of CV events

There were 18,148 participants (11.8%) in the total cohort with the primary endpoint of composite CV event or CV death during a median of 13 years of follow-up. In Table 3, the multivariable associations between geographic region, rural-urban residence, insurance status, and annual household income and the composite of CV events and CVD death are shown. There was no difference, after multivariable adjustment, in the association of geographic region with the primary endpoint of composite CV event or CV mortality when comparing participants in the Midwest (HR 0.97, 95% CI 0.92-1.02), West (HR 1.01, 95% CI: 0.97-1.06), or South (HR 1.02, 95% CI 0.98-1.08) to those in the Northeast in the total cohort. However, there was a significant interaction by BC status for geographic region and the composite endpoint (p-interaction=0.009). For example, for participants with BC, individuals in the South had the lowest risk of the composite outcome (HR 0.68, 95% CI 0.54-0.87) as compared to those in the Northeast. For individuals with BC, comparing those in the Midwest or the West with the Northeast was not significant. For individuals without BC, there was no difference in the composite outcome by geographic region.

**Table 3.**
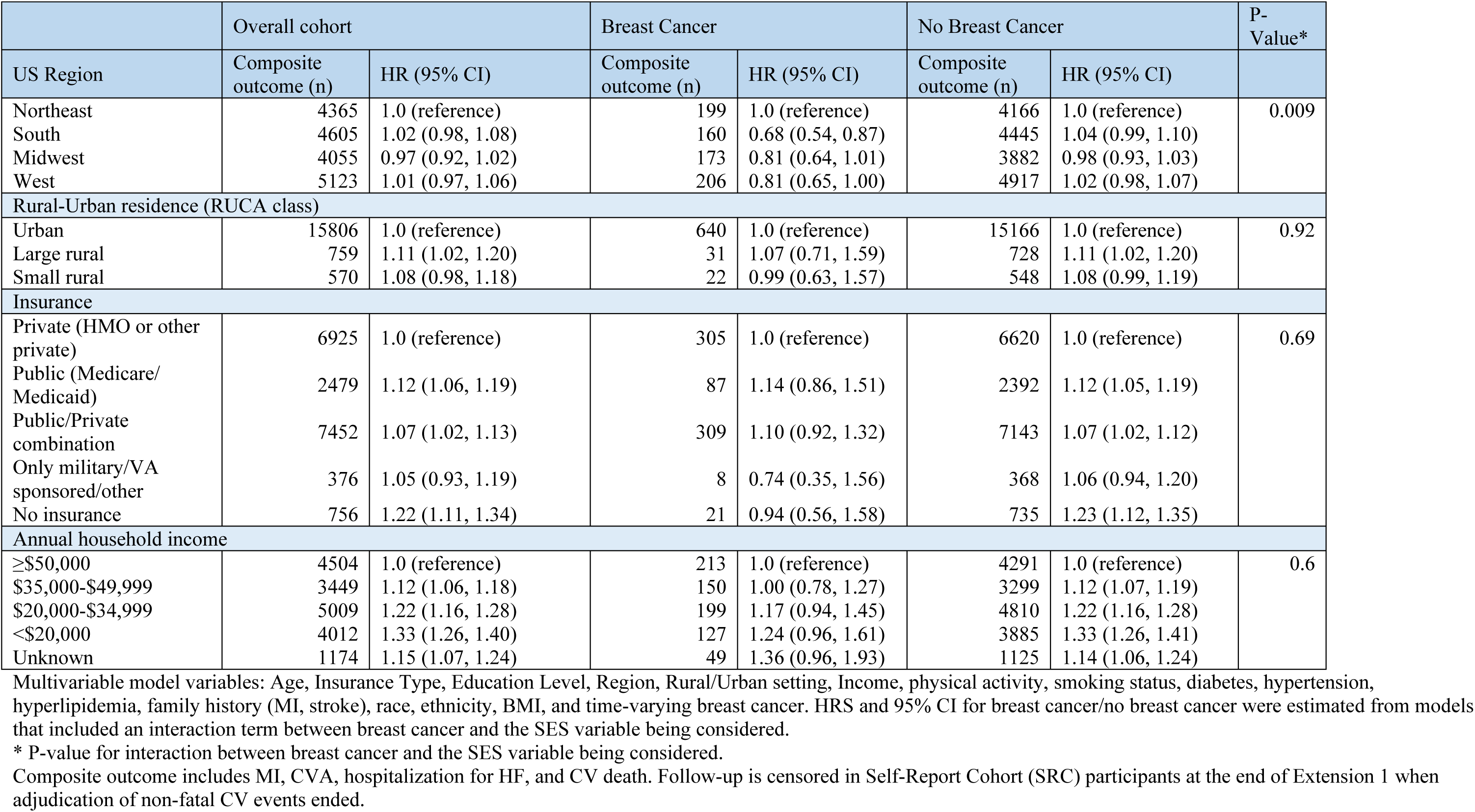
Association of Primary Endpoint (Composite CV event or CV mortality) with SES factors in Multivariable-Adjusted Analysis, overall and by incident breast cancer during follow-up.

|  | Overall cohort |  | Breast Cancer |  | No Breast Cancer |  | P-Value* |
| --- | --- | --- | --- | --- | --- | --- | --- |
| US Region | Composite outcome (n) | HR (95% CI) | Composite outcome (n) | HR (95% CI) | Composite outcome (n) | HR (95% CI) |  |
| Northeast | 4365 | 1.0 (reference) | 199 | 1.0 (reference) | 4166 | 1.0 (reference) | 0.009 |
| South | 4605 | 1.02 (0.98, 1.08) | 160 | 0.68 (0.54, 0.87) | 4445 | 1.04 (0.99, 1.10) |  |
| Midwest | 4055 | 0.97 (0.92, 1.02) | 173 | 0.81 (0.64, 1.01) | 3882 | 0.98 (0.93, 1.03) |  |
| West | 5123 | 1.01 (0.97, 1.06) | 206 | 0.81 (0.65, 1.00) | 4917 | 1.02 (0.98, 1.07) |  |
| Rural-Urban residence (RUCA class) |  |  |  |  |  |  |  |
| Urban | 15806 | 1.0 (reference) | 640 | 1.0 (reference) | 15166 | 1.0 (reference) | 0.92 |
| Large rural | 759 | 1.11 (1.02, 1.20) | 31 | 1.07 (0.71, 1.59) | 728 | 1.11 (1.02, 1.20) |  |
| Small rural | 570 | 1.08 (0.98, 1.18) | 22 | 0.99 (0.63, 1.57) | 548 | 1.08 (0.99, 1.19) |  |
| Insurance |  |  |  |  |  |  |  |
| Private (HMO or other private) | 6925 | 1.0 (reference) | 305 | 1.0 (reference) | 6620 | 1.0 (reference) | 0.69 |
| Public (Medicare/Medicaid) | 2479 | 1.12 (1.06, 1.19) | 87 | 1.14 (0.86, 1.51) | 2392 | 1.12 (1.05, 1.19) |  |
| Public/Private combination | 7452 | 1.07 (1.02, 1.13) | 309 | 1.10 (0.92, 1.32) | 7143 | 1.07 (1.02, 1.12) |  |
| Only military/VA sponsored/other | 376 | 1.05 (0.93, 1.19) | 8 | 0.74 (0.35, 1.56) | 368 | 1.06 (0.94, 1.20) |  |
| No insurance | 756 | 1.22 (1.11, 1.34) | 21 | 0.94 (0.56, 1.58) | 735 | 1.23 (1.12, 1.35) |  |
| Annual household income |  |  |  |  |  |  |  |
| ≥\$50,000 | 4504 | 1.0 (reference) | 213 | 1.0 (reference) | 4291 | 1.0 (reference) | 0.6 |
| \$35,000-\$49,999 | 3449 | 1.12 (1.06, 1.18) | 150 | 1.00 (0.78, 1.27) | 3299 | 1.12 (1.07, 1.19) | |
| \$20,000-\$34,999 | 5009 | 1.22 (1.16, 1.28) | 199 | 1.17 (0.94, 1.45) | 4810 | 1.22 (1.16, 1.28) | |
| <\$20,000 | 4012 | 1.33 (1.26, 1.40) | 127 | 1.24 (0.96, 1.61) | 3885 | 1.33 (1.26, 1.41) | |
| Unknown | 1174 | 1.15 (1.07, 1.24) | 49 | 1.36 (0.96, 1.93) | 1125 | 1.14 (1.06, 1.24) |  |

Multivariable model variables: Age, Insurance Type, Education Level, Region, Rural/Urban setting, Income, physical activity, smoking status, diabetes, hypertension, hyperlipidemia, family history (MI, stroke), race, ethnicity, BMI, and time-varying breast cancer. HRS and 95% CI for breast cancer/no breast cancer were estimated from models that included an interaction term between breast cancer and the SES variable being considered.
\* P-value for interaction between breast cancer and the SES variable being considered.
Composite outcome includes MI, CVA, hospitalization for HF, and CV death. Follow-up is censored in Self-Report Cohort (SRC) participants at the end of Extension 1 when adjudication of non-fatal CV events ended.

In the total cohort of women, rurality, non-private insurance status or no insurance, and household incomes <$50,000 were each associated with higher risk of CVD. In detail, for rurality (RUCA), participants in large rural areas (HR 1.11, 95% CI: 1.02-1.20) had higher risk of CV events or death compared to those in urban areas. Participants with public (Medicare/Medicaid) (HR 1.12, 95% CI 1.06-1.19), public/private combination insurance (HR 1.07, 95% CI 1.02-1.13) or no insurance (HR 1.22, 95% CI 1.11-1.34) had higher risk of CVD compared to those with private insurance. Categories of lower annual household income were linearly associated with increased risk of the composite CV outcome: $35,000-49,999 (HR 1.12, 95% CI 1.06-1.18), $20,000-34,999 (HR 1.22, 95% CI: 1.16-1.28), and <$20,000 (HR 1.33, 95% CI 1.26-1.40) compared to those in the reference group of ≥$50,000 (p-trend across categories=0.0002). However, we observed no effect modification by BC status for RUCA status, insurance, or household income (p = 0.92, p=0.69, and p=0.60, respectively).

### Risk of Mortality

As secondary endpoints, we evaluated CV mortality, BC mortality, and all-cause mortality in relation to geographic and (Socioeconomic status) SES factors (Table 4). For CV mortality in US regions, the South (HR 1.08 95% CI 1.02-1.14) and West (HR 1.13 95% CI 1.08-1.19) had higher CV mortality risk compared to the Northeast. Similarly, the South (HR 1.10, 95% CI 1.06-1.13) and West (HR 1.06, 95% CI 1.03-1.09) had an increased all-cause mortality risk compared to the Northeast. In contrast, the Midwest (HR 1.23, 95% CI 1.04-1.45), and South (HR 1.18, 95% CI 1.00-1.39) had a higher BC mortality risk compared to the Northeast but no significant BC mortality differences were found when comparing the West (HR 1.16, 95% CI 0.99-1.36) to the Northeast. Based on rural status, no significant differences in risk were observed between large rural and small rural areas compared to urban areas for CV mortality, BC mortality, or all-cause mortality (Table 4). When examining insurance status, we found that compared to private insurance, all other forms of insurance were associated with an increased risk of CV mortality and all-cause mortality. However, insurance status was not associated with BC mortality. When assessing household income and CV mortality, each income bracket below ≤$50,000 was associated with a higher risk of CV mortality and all-cause mortality. In contrast, when assessing household income and BC mortality, aside from a higher risk in individuals with an income < $20,000 (HR 1.26, 95% CI 1.04-1.53), no mortality differences were observed across other income brackets.

**Table 4.** Cause-specific mortality: CV, Breast cancer, other cause, and all-cause Mortality in relation to geographic and SES factors in the Overall Cohort.

|  | CV death |  | Breast Cancer Death |  | Other Death |  | All Death |  |
| --- | --- | --- | --- | --- | --- | --- | --- | --- |
| US Region | N | HR (95% CI) | N | HR (95% CI) | N | HR (95% CI) | N | HR (95% CI) |
| Northeast | 3518 | 1.0 (reference) | 343 | 1.0 (reference) | 7936 | 1.0 (reference) | 11797 | 1.0 (reference) |
| South | 3294 | 1.08 (1.02, 1.14) | 382 | 1.18 (1.00, 1.39) | 7578 | 1.10 (1.06, 1.14) | 11254 | 1.10 (1.06, 1.13) |
| Midwest | 3087 | 0.97 (0.92, 1.02) | 379 | 1.23 (1.04, 1.45) | 7182 | 1.03 (0.99, 1.06) | 10648 | 1.02 (0.99, 1.05) |
| West | 4545 | 1.13 (1.08, 1.19) | 449 | 1.16 (0.99, 1.36) | 9398 | 1.02 (0.99, 1.06) | 14392 | 1.06 (1.03, 1.09) |
| Rural-Urban residence (RUCA class) |  |  |  |  |  |  |  |  |
| Urban | 12733 | 1.0 (reference) | 1375 | 1.0 (reference) | 28242 | 1.0 (reference) | 42350 | 1.0 (reference) |
| Large rural | 568 | 1.04 (0.95, 1.14) | 47 | 0.77 (0.56, 1.06) | 1224 | 0.97 (0.91, 1.04) | 1839 | 0.98 (0.93, 1.04) |
| Small/isolated rural | 396 | 0.96 (0.86, 1.08) | 32 | 0.69 (0.47, 1.01) | 925 | 1.02 (0.95, 1.09) | 1353 | 0.99 (0.93, 1.05) |
| Insurance |  |  |  |  |  |  |  |  |
| Private (HMO or other private) | 4835 | 1.0 (reference) | 825 | 1.0 (reference) | 13624 | 1.0 (reference) | 19284 | 1.0 (reference) |
| Public (Medicare/Medicaid) | 2067 | 1.20 (1.13, 1.29) | 141 | 1.03 (0.81, 1.29) | 3783 | 1.14 (1.08, 1.19) | 5991 | 1.15 (1.11, 1.19) |
| Public/Private combination | 6784 | 1.14 (1.08, 1.20) | 485 | 1.07 (0.91, 1.27) | 12695 | 1.10 (1.06, 1.13) | 19964 | 1.10 (1.07, 1.13) |
| Only military/VA sponsored/other | 260 | 1.21 (1.05, 1.39) | 44 | 1.29 (0.92, 1.82) | 662 | 1.12 (1.02, 1.23) | 966 | 1.15 (1.07, 1.24) |
| No insurance | 393 | 1.47 (1.31, 1.67) | 51 | 0.78 (0.54, 1.12) | 1079 | 1.23 (1.14, 1.33) | 1523 | 1.27 (1.19, 1.35) |
| Annual household income |  |  |  |  |  |  |  |  |
| ≥\$50,000 | 3861 | 1.0 (reference) | 550 | 1.0 (reference) | 10078 | 1.0 (reference) | 14489 | 1.0 (reference) |
| \$35,000-\$49,999 | 2822 | 1.12 (1.06, 1.18) | 318 | 1.07 (0.92, 1.25) | 6426 | 1.08 (1.04, 1.12) | 9566 | 1.09 (1.06, 1.12) |
| \$20,000-\$34,999 | 3994 | 1.25 (1.18, 1.32) | 350 | 1.05 (0.89, 1.23) | 8275 | 1.16 (1.12, 1.20) | 12619 | 1.18 (1.14, 1.21) |
| <\$20,000 | 2836 | 1.39 (1.31, 1.48) | 236 | 1.26 (1.04, 1.53) | 5419 | 1.28 (1.23, 1.34) | 8491 | 1.31 (1.27, 1.36) |
| Unknown | 931 | 1.20 (1.10, 1.31) | 99 | 1.16 (0.90, 1.49) | 1896 | 1.09 (1.03, 1.16) | 2926 | 1.12 (1.07, 1.18) |

Multivariable model variables: Age, Insurance Type, Education Level, Region, Rural/Urban setting, Income, physical activity, smoking status, diabetes, hypertension, hyperlipidemia, family history (MI, stroke), race, ethnicity, and BMI.

## Discussion

To our knowledge, this is the largest study to evaluate how geography and the SDOH are associated with CV risk in both women with and without BC. Our findings can be summarized as follows: (1) We observed no difference in the composite endpoint of incident CV events or CV mortality in the total cohort by large geographic regions; however, there appeared to be effect modification by BC status. For individuals without BC, geographic region was not associated with the composite endpoint. However, women with BC living in the South appeared to have decreased risk of primary endpoint of composite CV event or CV mortality compared to those in the Northeast. (2) The risk for BC mortality was higher in the South and Midwest as compared to the Northeast. These findings underscore the importance of considering competing risk as the increased risk for other causes of mortality may impact accrued exposure time for CV events. (3) Independent of geographic region, the SDOH which included rurality, socioeconomic status, and insurance status appear to drive the increased risk for CV mortality. Participants in rural areas had higher CV risk compared to those in urban areas, those with public insurance had higher risk compared to those with private insurance, and those with lower household income had higher CV risk than those with higher income with a stepwise increased hazard by income strata. For RUCA, insurance status, and household income, there was no effect modification by BC status suggesting that individuals with BC are as vulnerable when they are socioeconomically disadvantaged. (4) Our findings suggest that regardless of geographic region, similar SDOH drive individual CV risk regardless of BC status.

In the US general population, the highest CV mortality rates are concentrated across Southern states, such as Mississippi, Kentucky, Oklahoma, Tennessee, Arkansas, Louisiana, Georgia, Texas, Alabama, and Missouri.^24^ Geographic concentrations in CVD risk have also shifted over time. A spatiotemporal study from 1873 to 2010 observed an overall CV mortality shift from Northeast to the South, with higher CV hospitalizations in Oklahoma, Kentucky, and West Virginia.^25^ For women, CV disease burden increased in Indiana, Kentucky, Michigan, Mississippi, Missouri, New Mexico, and South Dakota from 2010 to 2016.^26^ Multiple factors are responsible for these trends, including the higher prevalence of CV risk factors across Southern states as well as socioeconomic, healthcare environment, and health status factors.^27,28^ In our study, there was no difference across the composite CV event or CV mortality when comparing participants in the Midwest or West to those in the Northeast. Additionally, individuals with BC living in the South had a lower association of CV mortality risk compared with the Northeast. However, all-cause mortality, BC mortality, as well as CV mortality was found to be higher in the South compared to the Northeast similar to that seen in other studies.^26^ From our analysis, the higher BC mortality rates observed in the South may offer a possible explanation for why women with BC had decreased CV event risk, as the increased risk for BC mortality may impact accrued exposure time for CV events.

Differences between our study and prior large scale geographic studies may be attributed to differences in the WHI participant population and surveillance methods. The WHI enrolled healthy postmenopausal women volunteers from 40 major academic centers and may not reflect the general population. Additionally, the WHI surveillance methods did not include small-area surveillance data (i.e., county).^29^ We believe this should be highlighted, given that important CVD variations can be observed at the county-level.^30,31^ County-level data represent complex environments with variable distribution of community-level risk factors. Consequently, our lack of regional geographic differences across women with BC may be due to variations accounted for in a smaller scale including county-level data or rural-urban status.^32^ ^2930,3132^When stratified by rural/urban status, participants in rural areas had higher risk of CV events compared to those in urban areas. BC status did not confer increased CV mortality, but may be due to the relatively smaller number of women in BC located in large rural and small rural areas, resulting in a small sample size and null findings. When examining rural/urban status for BC mortality and all-cause mortality, no differences were found. Our finding is consistent with similar studies which observed that CV mortality rate was significantly higher in rural areas compared to urban locations.^33^ Rural regions have variations in access for the mentioned social factors which may contribute to adverse CV outcomes.^34^ Rural regions tend to have substandard housing, more limited transportation options, lower food access, and lower rates of health literacy, all which have been implicated in poorer CV outcomes.^34–38^ In a study of BC survivors, the most rural counties had a 31% increased CV mortality risk compared with the most urban counties.^32^ Rural areas may significantly contribute to these outcomes, given limited access to health care compared to urban areas. One study identified travel distance to treatment centers as a substantial barrier to women with BC in rural areas.^39^

Independent of geographic region and BC status, we observed that SDOH drive an increased risk for CV events and CV mortality in our cohort. While the stratified results for those with BC did not reach significance, the point estimates were generally similar, and the lack of significance may have been driven by the limited sample size for the BC-only cohort. Therefore, it is possible that the stratified analyses for BC status may be underpowered. However, the lack of effect modification of the SDOH by BC status suggests that individuals with BC remain a vulnerable population even though women with cancer may be more likely to be established in the medical system for their oncologic care. Beyond CV mortality, we found that SDOH, including insurance status and household income, was associated with an increased risk for all-cause mortality. The SDOH encompasses the characterization of access to medical care in regards to neighborhood quality, housing, education, healthcare, transportation, and food, and has been implicated by others in adverse CV outcomes.^40^ SDOH has been associated with worse cardiovascular health in adult cancer survivors in the U.S.^41^ Across women with BC, lack of private insurance and low socioeconomic status has been associated with a higher CV mortality.^42–44^ In a recent analysis, CV mortality was 41% higher among women with BC in the counties with the lowest SES compared with the wealthiest counties.^32^ Future studies should explore the association between SDOH and mortality to further examine possible drivers of these disparities.

In our study, we emphasize the impact of socioeconomic factors on CV risk in both women with and without BC. The increased risk of SDOH on adverse CV outcomes may be explained by lower SES, financial barriers, and limited access to medical care. For women with BC, BC clinical guidelines outline CV screening and monitoring before, during and after receipt of cardiotoxic cancer treatment. However, women with BC lacking insurance or with lower financial resources may receive less medical care and attention and access, despite complex survivorship needs.^45^ The findings in our study underscore the need for patient-centered approaches that consider the interplay between social and economic factors to optimize CV outcomes in women with poor SDOH. Further research is warranted to elucidate the mechanisms underlying the observed differences in CV risk and to develop targeted interventions for women with and without BC.

### Limitations

This study has several limitations. The WHI cohort is predominantly White, age 50-79 years, and participants were primarily recruited from major academic health centers in 24 states and the District of Columbia.^29,46^ Although recruitment areas included rural, suburban, and rural populations, rural areas included fewer women compared to larger density population areas. In addition, SDOH factors, such as income levels and insurance status, may have changed to varying degrees over time and follow up period, especially as participants entered retirement. However, despite this, one major strength of this study is that we demonstrated in the WHI population, that poor socioeconomic status, defined by lower income, public insurance, as well as rurality are all associated with health outcomes, an important finding that can impact individuals in all sectors of the country. Further research is needed to further evaluate the long-term association between SES, CV risk, and cancer across the United States.

## Conclusions

In this population of postmenopausal women with and without BC, we found that CV risk and mortality is highest in individuals in rural areas, from lower income status and with public or no insurance regardless of geographic region. Further studies are needed to evaluate population-level interventions to improve CV outcomes among underserved women with BC. Future public health initiatives need to account for these disparities in communities across the country.

## Data Availability

Data are accessible through the established data sharing policies for the Women?s Health Initiative at https://www.whi.org/page/working-with-whi-data.

## Acknowledgements

The authors would like to thank the WHI investigators and staff for their dedication, and the study participants.

---

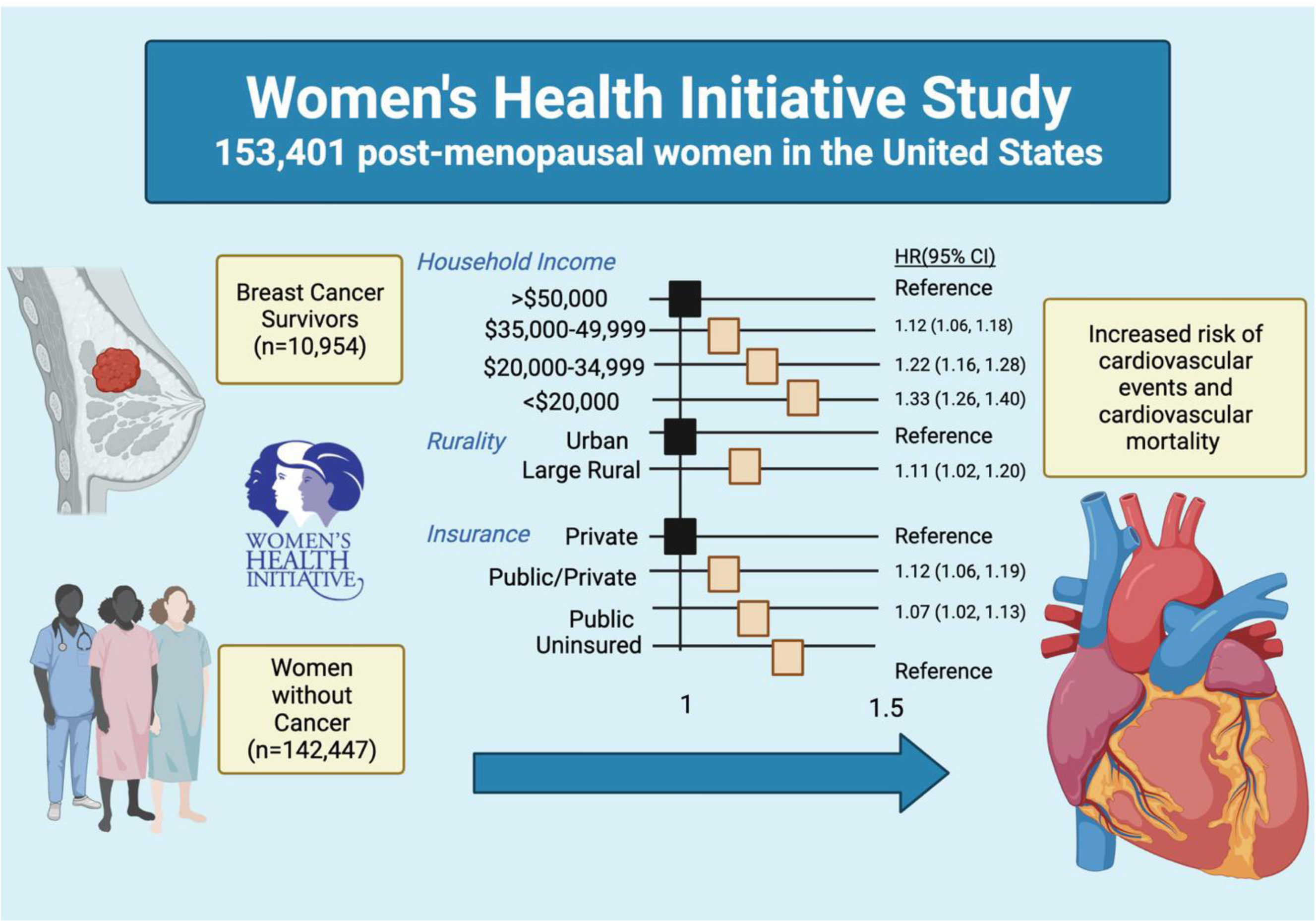
Through the Women’s Health Initiative study, 153,401 post-menopausal women in the United States were evaluated for CVD risk stratified by geography, rural status, and social determinants of health. Risk factors associated with increased risk for cardiovascular events and cardiovascular mortality included having an annual income less than $50,000, rurality, and having non-private insurance.

